# Classification and management strategy of spontaneous carotid artery dissection

**DOI:** 10.1101/2023.10.23.23297442

**Authors:** Baoning Zhou, Chong Li, Zhouyang Jiao, Hui Cao, Peng Xu, Shirui Liu, Zhen Li, Zhaohui Hua

**Affiliations:** Department of Endovascular Surgery, The First Affiliated Hospital of Zhengzhou University, Zhengzhou, China; The Vascular Care Group, Darien, CT, America

**Keywords:** Carotid artery, dissection, Ischemic stroke, Stenting, Endovascular treatment

## Abstract

**Background and Purpose:** Spontaneous carotid artery dissections (sCAD) are the main cause of stroke in middle-aged and young people. There is still a lack of clinical classification to guide the management of sCAD. We reviewed our experience with 179 sCAD patients and proposed a new classification for sCAD with prognostic and therapeutic significance.

**Methods:** This is a retrospective review of prospectively collected data from June 2018 to June 2023 of sCAD patients treated at a large tertiary academic institution in an urban city in China. Depending on the degree of luminal narrowing and pseudoaneurysm formation on imaging, sCAD was classified into four types. Type IV dissections were divided into type IVA and type IVB dissections according to the presence of intracranial occlusion.

**Results:** A total of 179 patients and 197 dissected arteries met the inclusion criteria. More than two-thirds of type I dissections are completely recanalized after antithrombotic therapy, and only one case (1.8%) had recurrent ischemic stroke. A total of 38 % of type II dissections and 73% of type III dissections received endovascular treatment (EVT) for persistent flow-limited dissections, enlargement of pseudoaneurysm, or aggravation of clinical symptoms despite antithrombotic therapy. Type IV dissections are more likely to lead to the occurrence of ischemic stroke, and presented with more severe symptoms. About 33% of type IVB dissections received emergent intervention due to intracranial occlusion or aggravation of symptoms after medical treatment.

**Conclusions:** This study proposes a novel and more comprehensive classification method and management strategy for sCAD. Antithrombotic therapy is beneficial to reduce the risk of recurrent stroke for stable sCAD like type I. Non-emergent EVT can be an alternative therapeutic approach for patients who meet indications as in type II to IVA. Urgent procedure with neurovascular intervention might be needed in those with type IVB SCAD. The short-term results of EVT for sCAD are encouraging, and long-term device-related and functional outcome should be elucidated.

---

Carotid artery dissections (CAD) are a rare cause of stroke, accounting for about 2.5% of all stroke patients^1^, but it is the main cause of stroke in middle-aged and young people, which accounts for 8-25% of the causes of ischemic stroke in people under 45 years old^2, 3^. More than 90% of CAD is spontaneous carotid artery dissections (sCAD), which occur spontaneously or are secondary to minor trauma, such as neck massage, weightlifting, yoga, and severe cough^4^. Extracranial sCAD is more common than intracranial sCAD, because of the increased mobility, longer anatomical distance, and vulnerability to mechanical stress^5, 6^. Since most carotid artery dissection heals spontaneously after antithrombotic therapy, European and American clinical guidelines recommend it as the initial choice for sCAD patients’ management^7, 8^. Perry and colleagues proposed the Borgess classification of sCAD in 2013, which helps to plan medical treatment and predict healing following medical treatment. Endovascular treatment (EVT) for patients with sCAD has also been proven to be safe and effective in recent years^9-11^. Nevertheless, there is no clinical classification to guide the interventional or surgical management of sCAD. We retrospectively analyzed the clinical and imaging findings of sCAD patients admitted to the First Affiliated Hospital of Zhengzhou University in the past 5 years, throughout the patient’s clinical course, and evolution on follow-up, to develop a new classification with prognostic and therapeutic significance.

## Methods

This study was approved by the ethical committee and Institutional Review Board of the First Affiliated Hospital of Zhengzhou University (Henan, China), and rules according to the Declaration of Helsinki were followed. All patients treated for sCAD in the First Affiliated Hospital of Zhengzhou University from June 2018 to June 2023 were prospectively collected and retrospectively analyzed. We included all patients diagnosed as sCAD by CTA, MRA, or DSA. Exclusion criteria were pure intracranial dissection, aortic dissection with extension to carotid arteries, and non-spontaneous CAD caused by severe trauma or iatrogenic operation. The demographic data, procedural, symptoms upon presentation, medical comorbidities, baseline stroke severity measured by the National Institutes of Health Stroke Scale (NIHSS), and degree of stenosis. Clinical follow-up was ascertained from outpatient or inpatient medical record review and imaging, including revascularization condition, modified Rankin scale (mRS) score at 90 days, recurrent cerebral ischemic events, and symptomatic intracranial hemorrhage (SICH).

### Classification of sCAD

Based on the large single center cohort and imaging findings, we developed a new classification for sCAD (Figure 1). Type I and type II dissections were defined according to whether the luminal narrowing caused by dissection or intramural hematoma was less than 70%. The degree of stenosis was estimated by comparing the diameter of the dissection site with the diameter of the normal site at the proximal end of the dissection. If the proximal dissection is located at the level of the carotid bulb, the contralateral normal artery is used as the comparison. Type III dissections were defined as the formation of dissecting pseudoaneurysms. Complete occlusion of the lumen was defined as type IV dissections divided into two subtypes. Type IVA dissections were defined as the occlusion segment confined to the extracranial carotid artery. Tandem occlusions were classified as type IVB dissections, which was described as occlusion of both extracranial and intracranial arteries^6^.

**Figure 1.**
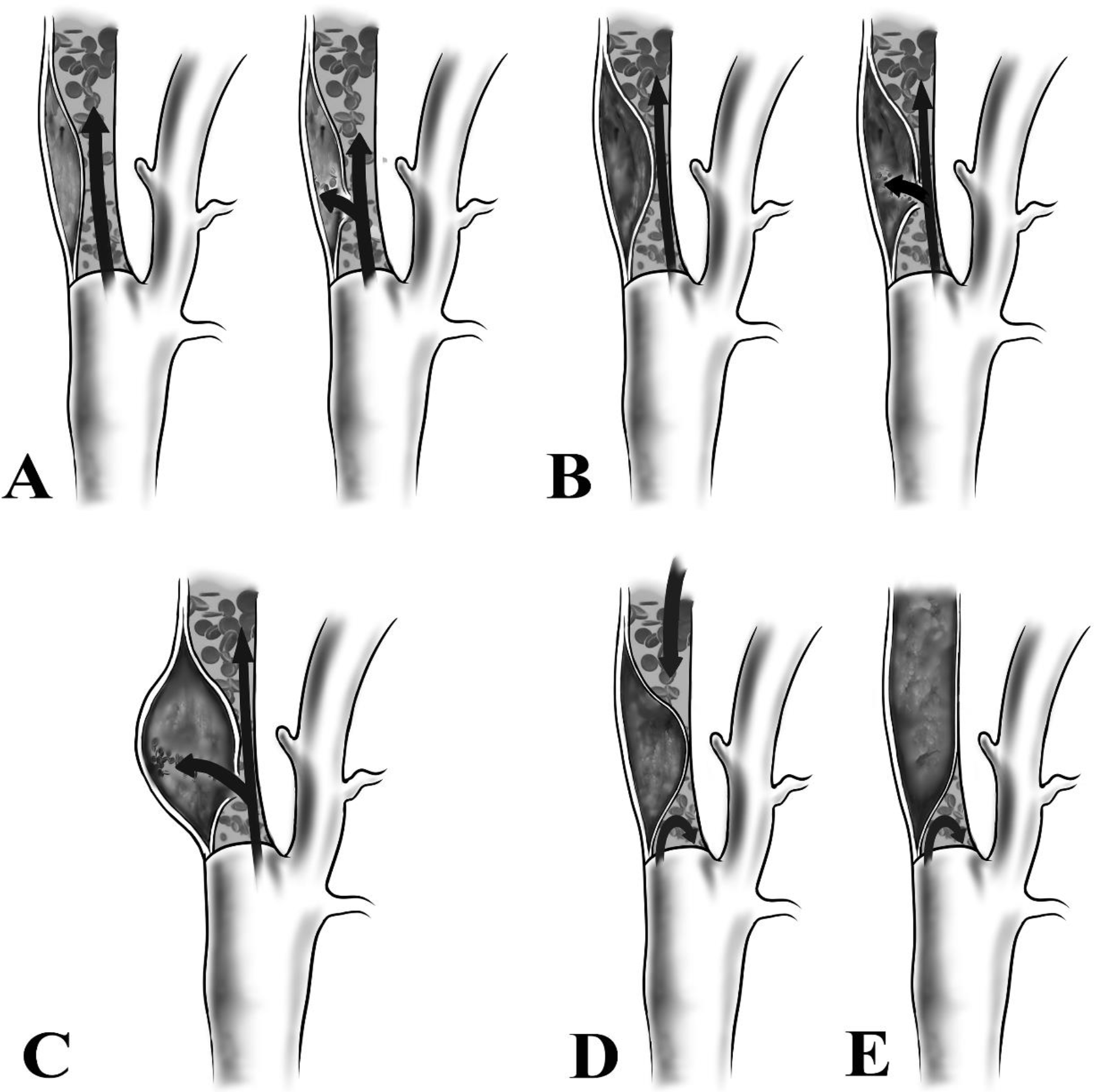
Classification of spontaneous carotid artery dissections on computed tomography angiography volume rendered images: (A) Type I, intramural hematoma or dissection with < 70% luminal narrowing; B: Type II, intramural hematoma or dissection with ≥ 70% luminal narrowing; C: Type III, dissecting pseudoaneurysm; D: Type IVA, extracranial carotid artery occlusion; E: Type IVB, Tandem occlusion.

### Patients and methods

Antithrombotic therapy is still the initial choice for patients with sCAD. All patients with sCAD were immediately given anticoagulant (a therapeutic dose of low-molecular-weight heparin) or antiplatelet (aspirin and clopidogrel in dual combination) therapy when there was no antithrombotic contraindication. Patients with indications for thrombolytic therapy^7^ received intravenous thrombolysis with rt-PA within 4.5 hours after the onset of acute stroke. EVT was recommended when the patient presented with the following indications: patients with recurrent cerebral ischemic events, persistent symptoms, continuous enlargement of dissecting pseudoaneurysms, or persistent flow-limiting dissection after antithrombotic therapy.

### Definitions

We defined sCAD by radiological criteria as the presence of a double lumen, intimal flap, long tapered arterial stenosis, focal fusiform or blister-like dilatation, flame-shaped occlusion, or intramural hematoma (hyperintense signal on fat-saturated T1-weighted sequence) on neuroimaging (computed tomography angiography (CTA), magnetic resonance imaging or magnetic resonance angiography (MRA), and/or digital subtraction angiography (DSA)^6^. Local signs and symptoms included neck pain, headache, Horner’s syndrome, and retinal infarction. Patients with both ischemic and local symptoms were classified under ischemic symptoms. During hospitalization, the patient has acute events such as thromboembolism, and the symptoms persist or aggravate after medical treatment, to receive endovascular or surgical treatment, which is defined as acute phase intervention. The reconstruction of the vascular lumen to normal anatomy in vascular imaging is defined as healing. Extracranial or intracranial (i.e. carotid artery to the furthest intracranial segments) occlusion as absent contrast medium filling of the examined arterial segment^12^. Excellent outcome defined as mRS 0–2 at 90-day follow-up.

### Statistical analysis

SPSS version 26.0 (SPSS Inc., Chicago, Illinois, United States) was used for statistical analysis of the data. Continuous variables were expressed as mean ± standard deviation and comparisons were made using the independent samples t-test; categorical variables were expressed as counts (percentages) and differences between two or more groups were analyzed using the χ2 test or Fisher’s Exact test, and the Mann–Whitney U test for continuous variables. Statistical significance was defined as a p-value < .05 two-sided).

## Results

### Patient characteristics

Among 202 cases of CAD during the observation period, 179 patients and 197 dissected arteries met the inclusion criteria. We excluded 19 patients with pure intracranial artery dissections and 4 patients caused by severe trauma. The mean age of the 179 sCAD patients was 49.5 years, 78% were male, 18 patients (10%) had bilateral sCAD, and the mean duration of follow-up was 26 months (range 3–59 months). Cerebral ischemic events including ischemic stroke (89 cases, 49.7%) and transient ischemic attack (TIA) (27 cases, 15.4%) were the main clinical presentations of sCAD patients. Hypertension (50.3%) and hyperlipidemia (23.5%) were the most common comorbidities in patients with sCAD. Thirty-seven (20.7%) patients indicated for thrombolysis received intravenous thrombolytic therapy. Five (2.8%) patients were not treated with anticoagulation or antiplatelet therapy due to the presence of subarachnoid hemorrhage.

According to our classification, there were 56 (28.4%) type I, 50 (25.4%) type II, 60 (30.5%) type III, and 31 (15.7%) type IV dissections. We compared baseline characteristics and prognosis among the four groups. Baseline characteristics are shown in Table 2. No statistically significant difference was found in sex ratio, BMI, sides, risk factors, thrombolytic therapy ratio, and hospitalization length among the four types. The mean age in type I is older than in the other three types (group-to-group comparison, *p*<.05). The proportion of ischemic stroke, NIHSS score at admission, NIHSS score at discharge, and mRS at discharge were significantly higher in type IV than in type I, and type III (group-to-group comparison, *p*<.001), and NIHSS score at discharge were significantly higher in type IV than in type II (*p*<.05). Thus, type IV dissections are more likely to lead to the occurrence of ischemic stroke, and the symptoms of type IV dissections are more serious.

**Table 1.** Baseline Characteristics.

| Characteristics | Type I (n=56<sup>*</sup>) | Type II (n=50<sup>*</sup>) | Type III (n=60<sup>*</sup>) | Type IV (n=31<sup>*</sup>) | p value |
| --- | --- | --- | --- | --- | --- |
| Male | 43(77) | 36(72) | 50(83) | 27(87) | .312 |
| Age, mean (SD), y | 53.5 ± 11.5 | 46.4 ± 10.9 | 48.9 ± 9.2 | 47.0 ± 11.5 | .004 |
| BMI, mean (SD) | 24.4 ± 3.4 | 24.1 ± 4.3 | 24.8 ± 2.8 | 24.1 ± 3.6 | .766 |
| Initial NIHSS, median(IQR) | 0(0-2.00) | 3(0.30-5.80) | 2(0-3.30) | 4(2.00-6.50) | <.001 |
| NIHSS at discharge, median(IQR) | 0(0-1.00) | 0(0-2.00) | 0(0-1.00) | 2(0-3.00) | <.001 |
| Presentation |  |  |  |  | <.001 |
| Ischemic stroke | 16(29) | 32(64) | 26(43) | 22(71) |  |
| Local signs and symptoms | 11(20) | 9(18) | 10(17) | 4(13) |  |
| TIA | 16(29) | 9(18) | 13(22) | 5(16) |  |
| Asymptomatic | 13(23) | 0 | 11(18) | 0 |  |
| Site of CAD |  |  |  |  | .227 |
| Left | 23(41) | 29(58) | 34(57) | 18(58) |  |
| Right | 33(59) | 21(42) | 26(43) | 13(42) |  |
| Risk factors |  |  |  |  |  |
| Hypertension | 29(52) | 20(40) | 32 (53) | 16(52) | .508 |
| Dyslipidemia | 12(21) | 13(26) | 16 (27) | 7 (23) | .905 |
| Smoking | 13(23) | 9(18) | 13 (22) | 10 (32) | .517 |
| Diabetes mellitus | 7(13) | 4(8) | 9(15) | 6(19) | .494 |
| Carotid tortuosity | 6(11) | 5(10) | 4(7) | 1(3) | .592 |
| Neck massage | 6(11) | 1(2) | 7(12) | 0(0) | .062 |
| mRS at discharge (SD) | 0(0-0) | 0.5(0-1.00) | 0(0-1.00) | 1(0-2.00) | <.001 |
| Received stroke thrombolysis | 7(13) | 14(28) | 10(17) | 9(29) | .117 |
| Treatment |  |  |  |  | <.001 |
| Medication | 56(100) | 31(62) | 16(27) | 19(61) |  |
| Endovascular treatment | 0 | 19(38) | 43(72) | 11(36) |  |
| Surgery | 0 | 0 | 1(2) | 1(3) |  |
| Anti-thrombotic type selection |  |  |  |  | <.001 |
| Anticoagulants | 6(11) | 8(16) | 3(5) | 12(39) |  |
| Antiplatelets | 46(82) | 42(84) | 55(92) | 19(61) |  |
| Not Anti-thrombotic | 4(7) | 0 | 2(3) | 0 |  |
| Hospitalization length, median (IQR), d | 10(7.00-14.00) | 9(5.25-14.75) | 10(7.00, 14.00) | 11(8.00, 14.50) | .434 |

Data are presented as n (%), mean ± standard deviation, or median (Interquartile range). CAD = carotid artery dissection.
\* Represents the number of sCAD.

**Table 2.** Baseline Characteristics of Type IVA and Type IVB dissections.

| Table 2. Baseline Characteristics of Type IVA and Type IVB dissections |  |  |  |
| --- | --- | --- | --- |
| Characteristics | Type IV A (n=7<sup>*</sup>) | Type IV B (n=24<sup>*</sup>) | p value |
| Male | 7(100) | 20(83.3) | .55 |
| Age, mean (SD), y | 48.4±11.9 | 42.1±9.0 | .21 |
| Initial NIHSS, median (IQR) | 4(2.00-11.00) | 4(3.00-5.50) | .94 |
| NIHSS at discharge, median (IQR) | 2(0-3.00) | 1(1.00-2.00) | .74 |
| Presentation |  |  | 1 |
| Ischemic stroke | 5(71.4) | 17(70.8) |  |
| Head or neck pain | 1(14.3) | 3(12.5) |  |
| TIA | 1(14.3) | 4(16.7) |  |
| Site of CAD |  |  | .67 |
| Left | 5 (71.4) | 13(54.2) |  |
| Right | 2 (28.6) | 11 (41.7) |  |
| Site of intracranial occlusion |  |  |  |
| ICA | / | 16(66.7) |  |
| M1, M2 | / | 7(29.2) |  |
| A1 | / | 1(4.2) |  |
| mRS at discharge, median (IQR) | 1(1.00-2.00) | 2(0.50-2.50) | .80 |
| Received stroke thrombolysis | 1(14.3) | 4(16.7) | 1 |
| Treatment |  |  | 1 |
| Medication | 6(85.7) | 14(58.3) |  |
| Endovascular treatment | 1(14.3) | 9(37.5) |  |
| Surgery | 0 | 1(4.2) |  |
| Acute phase intervention | 0 | 8(33.3) | .15 |
| Complications after treatment |  |  | .30 |
| SICH | 0 | 1(4.2) |  |
| Death | 0 | 1(4.2) |  |
| Stent occlusion | 0 | 2(8.3) |  |
| Operation failure | 0 | 1(4.2) |  |
| Hospitalization length, median (IQR), d | 8.6±2.1 | 13.4±7.9 | .013 |

Data are presented as n (%), mean ± standard deviation, or median (Interquartile range). NIHSS = National Institutes of Health Stroke Scale. TIA= transient ischemic attack. CAD = carotid artery dissection. ICA= internal carotid dissection. SICH = symptomatic intracranial hemorrhage
\* Represents the number of sCAD.

Type IV dissections are divided into type IV A (7, 22.6%) and type IV B (24, 77.4%) according to whether there are tandem occlusions. Individual characteristics of type IVA and type IVB dissections are shown in Table 1. The hospitalization length was significantly higher in type IVB dissections than in type IVA dissections (8.6±2.1 versus 13.4±7.9, *p*<.05).

### Endovascular interventions and surgery

A total of 69 sCAD patients and 75 dissected arteries received endovascular or surgical treatment. Successful reconstruction at the end of the procedure was achieved in 73 (97.3%) dissected arteries. The overall complication rate was 6.7%. The incidence of complications in type IV dissections was significantly higher than that in type III dissections (*p*<.05). Six patients were treated by EVT for bilateral carotid arteries. Procedure-related information is shown in Table 2. Seventy-three (97.3%) dissected arteries were treated with EVT. Several endovascular techniques were used, including thrombectomy with stent-retriever (4, 5.3%), catheter thrombus aspiration (4, 5.3%), angioplasty (23, 30.7%), aneurysm coil embolization (8, 10.7%) and stenting (73, 97.3%). Fourteen (18.7%) patients received acute phase surgery due to distal embolization or aggravation of symptoms. All patients recovered well after intervention except one patient who received acute phase surgery for acute recurrent massive cerebral infarction died from brain herniation secondary to cerebral edema caused by intracranial hemorrhage. Two patients appeared with ischemic stroke or symptomatic intracranial hemorrhage after the operation and recovered well after conservative treatment. After endovascular recanalization in 2 patients with type IV dissections, asymptomatic stent occlusions were found 1 week after operation. One patient with a Type II dissection was unable to enter the true lumen due to the long-dissected segment and failed stent reconstruction. Two patients received open surgery. One patient with type III dissection underwent carotid endarterectomy and aneurysm resection. The other patient with type IVB dissection underwent open embolectomy but failed because of the long occlusion of intracranial carotid artery.

### Follow-Up Outcomes

Overall, the incidence of ischemic stroke and TIA was 3.9% and 2.8% during the follow-up period (mean, 26 months; range, 3–59 months). Excellent outcome was achieved in 168 (93.9%) patients. All cerebral ischemic events occurred in patients receiving conservative treatment. Two patients had asymptomatic mild in-stent stenosis 6 months after the operation. One patient was re-admitted for carotid artery stenting due to secondary dissection at the distal end of the stent 1 year after the endovascular intervention. The mRS at 90 days in type IV was significantly higher than in type I (*p*<.05). Table 4 reports the prognosis and the occurrence of complications during the follow-up period of patients in the four types of dissections. One patient with type I dissection died of severe pneumonia 19 months after discharge. The recanalization of the carotid artery receiving conservative treatment is shown in Table 5. Fifty (41.0%) sCAD was completely recanalized at 1 year with continuous antithrombotic therapy.

**Table 3.** Endovascular and Surgical Interventions of the sCAD Patients.

| Procedural Variables | Type II<br>(n=19*) | Type III (n=44*) | Type IV<br>(n=12*) | p value |
| --- | --- | --- | --- | --- |
| Indication for intervention |  |  |  | <.001 |
| Thromboembolism | 0 | 2(4.5) | 7(58.3) |  |
| Enlarging dissecting pseudoaneurysm | 0 | 14(31.8) | 0 |  |
| Symptoms persist or aggravate | 6(31.6) | 11(25.0) | 2(16.7) |  |
| Flow-limiting dissection | 13(68.4) | 17(38.6) | 3(25.0) |  |
| Treatment |  |  |  | <.001 |
| Stenting | 19(100) | 35(79.5) | 3(25.0) |  |
| Mechanical thrombectomy + stenting | 0 | 0 | 8(66.7) |  |
| Aneurysm coil embolization + stenting | 0 | 8(18.2) | 0 |  |
| Surgery | 0 | 1(2.3) | 1(8.3) |  |
| Failed surgery | 1(5.3) | 0 | 1(8.3) | .20 |
| Postoperative complications |  |  |  | <.001 |
| Ischemic stroke | 1(5.3) | 0 | 0 |  |
| SICH | 0 | 0 | 1(8.3) |  |
| Stent occlusion | 0 | 0 | 2(16.7) |  |
| Death | 0 | 0 | 1(8.3) |  |
| Adverse events during the follow-up period |  |  |  | .60 |
| In-stent stenosis | 1(5.3) | 0 | 1(8.3) |  |
| New entry | 0 | 1(2.3) | 0 |  |

Data are presented as n (%). sCAD= spontaneous carotid artery dissections. SICH = symptomatic intracranial hemorrhage

**Table 4.** Outcomes and Events During Follow-up.

| Outcome and Event | Type I (n=56*) | Type II (n=50*) | Type III (n=60*) | Type IV (n=30*) | p value |
| --- | --- | --- | --- | --- | --- |
| mRS at 90-day, median (IQR) | 0(0-0) | 0(0-1.00) | 0(0-1.00) | 0.5(0-2.00) | .033 |
| Cerebral ischemic events |  |  |  |  |  |
| Ischemic stroke | 1(1.8) | 3(6.0) | 2(3.3) | 1(3.3) | .92 |
| TIA | 0 | 0 | 3(5.0) | 0 | .13 |
| Death | 1(1.8) | 0 | 0 | 0 | .43 |
| Follow-up time, median (IQR) | 23.5(13.75-36.25) | 27(16.50-35.00) | 21(12.00-38.50) | 23(12.50-38.00) | .92 |

Data are presented as n (%) or median (Interquartile range). \* Represents the number of sCAD.
mRS = modified Rankin Scale. TIA= transient ischemic attack.

**Table 5.** Recanalization of patients received continuous antithrombotic therapy during follow-up at 1 year.

| Outcome and Event | Type I (n=56*) | Type II<br>(n=31*) | Type III<br>(n=16*) | Type IV<br>(n=19*) |
| --- | --- | --- | --- | --- |
| Recanalization | 29(51.8) | 19(61.3) | 0 | 2(10.5) |
| Partial recanalization | 27(48.2) | 12(38.7) | 16(100) | 12(63.2) |
| Occlusion | 0 | 0 | 0 | 5(26.3) |

Data are presented as n (%). \* Represents the number of sCAD.

## Discussion

Spontaneous carotid artery dissection (sCAD) is the most common cause of ischemic stroke in young and middle-aged patients, especially those without vascular risk factors^13, 14^. Intramural hematoma or pseudoaneurysm formed by arterial dissection can lead to stenosis or occlusion of the arterial lumen, resulting in distal thromboembolism, hypoperfusion, and cerebral ischemia. Currently, there is no accepted clinical classification to guide the management of sCAD. Perry *et al*.^15^ proposed the Borgess classification in 2013 and divided sCAD into type I and type II based on the presence of intimal tear in the carotid artery on imaging. The Borgess classification proved that dissections without intimal tear (type I) had a higher rate of healing than dissections with intimal tear (type II) after antithrombotic therapy, which guides the medical management of sCAD^15^. Nevertheless, with the wide application of EVT technology in the treatment of sCAD, a new classification is needed to guide the modern management of sCAD. We reviewed our experience over the past 5 years and the relevant literature and presented a new classification of sCAD with prognostic and therapeutic implications, using our large cohort of patients with sCAD.

Previous studies have shown that after receiving antithrombotic therapy, 33-90% of sCAD patients experienced improvement or complete recanalization of the true lumen within 6 months^16, 17^; dissecting pseudoaneurysms in 40-50% of patients could shrink or disappear but also can be progressively enlarged to compress the true lumen or cause embolism^18, 19^. The probability of recurrent cerebral ischemic events in patients with sCAD is 0-13%, and it is mainly concentrated in the first two weeks after onset^20^. Some studies believe that the main cause of ischemic stroke in sCAD patients is the distal embolism caused by thromboembolic shedding, rather than hypoperfusion^21, 22^. The prevention of the recurrent ischemic stroke caused by the embolism has become the key to the management of sCAD. Because of the high rate of self-healing and low recurrence of ischemic events in sCAD patients treated with antithrombotic therapy, the European Stroke Organization, the Society for Vascular Surgery, and the American Heart/Stroke Association guidelines for the management of extracranial carotid artery dissection all recommend antithrombotic therapy as the first option for the initial treatment of sCAD^8, 23, 24^. Anticoagulant and antiplatelet therapy are equally effective in preventing stroke recurrence and promoting vascular remodeling. In the CADISS and TREAD-CAD, two prospective, multicenter, RCT studies showed, there is no significant difference in prognosis and safety between anticoagulant and antiplatelet therapy in patients with CAD, including prevention of recurrent cerebral ischemic events, residual stenosis, and recanalization^25, 26^, which is the same as the results of our center. In our study, nearly half of the sCAD patients have initial symptoms of ischemic stroke. Furthermore, all recurrent ischemic events occurred in patients who had manifested ischemic events, whereas recurrent ischemic events were absent in patients with only local symptoms^25^. Therefore, the management of patients with initial symptoms of stroke should be more cautious. Until recently, there is no strong evidence to support the indications, efficacy, and safety of surgical or endovascular treatment in patients with sCAD after the failure of conservative treatment^2^. According to the 2011 guidelines for the management of extracranial carotid and vertebral artery diseases from the Society of Vascular Surgery, surgical or endovascular treatment may be considered after failure of conservative treatment, and EVT is generally considered better than open surgery^27^. EVT aims to improve distal perfusion and reduce stroke and death in sCAD patients by providing immediate revascularization with a high success rate and an acceptable post-operative complication rate, as demonstrated in previous studies^6, 9, 20, 28, 29^. However, EVT in acute phase is not recommended unless the patient’s symptoms deteriorate rapidly due to thromboembolism or hypoperfusion. The systematic review and meta-analysis by Bontinis *et al*. included 24 relevant trials with a total of 1224 patients with CAD who underwent carotid artery stenting (CAS)^11^. This study was divided into a group with immediate CAS and a group with CAS after the failure of medical treatment, according to the timing of the EVT. The results show that CAS after the failure of medical treatment has lower mortality and complication rates than immediate CAS, confirming that EVT should be used as a second-line choice after failure of antithrombotic therapy. Carotid stent selection is not supported by studies or data, and therefore cannot be recommended.

In our series, all type I dissections responded well after antithrombotic therapy and did not require further endovascular or surgical treatment. More than one-quarter of type I dissections have clinical manifestations of ischemic stroke, but the recurrence rate of ischemic events during follow-up was only 1.8% and meanwhile the recanalization rate of type I dissections after one year of antithrombotic therapy was 51.8%. Asif *et al*. observed carotid stenosis >70% is considered to have hemodynamic significance and a higher possibility of recurrence of cerebral ischemic events, which is consistent with our results^30^. For type II dissections, 19 (38%) dissected arteries received CAS after medical treatment for persistent flow-limiting dissections. Among the remaining 31 type II dissections receiving medical treatment, there were 3 (9.7%) cases of ischemic stroke likely caused by thrombus embolism during the follow-up period. As for type II dissections with non-healing stenosis of a severe degree stenosis (>70%) on follow-up imaging after medical treatment, EVT should be considered prevent clinical deterioration. A total of 96.4% and 94.0% of type I and type II dissections achieved excellent outcomes, respectively. This is in line with previous studies^25, 26^.

Type III dissections accounted for about 30% of all sCAD in this cohort, categorized as having dissecting pseudoaneurysm. Although a significant proportion of type I and type II dissections improved or healed with medical therapy, none of type III dissections healed. The formation of dissecting pseudoaneurysms increases the risk of thromboembolism or further enlargement with obliteration of the vessel, leading to ischemic stroke or aneurysm rupture. In our follow-up, recurrent stroke occurred in 6.7 % (4/60) of type III dissections during the follow-up period, and two of them received EVT. No rupture of type III dissections occurred during follow-up, consistent with previous studies^19, 31^. CAS is the most common method of EVT for type III dissections with the presence of persistent flow-limiting dissections, continued dissecting pseudoaneurysm enlargement, or persistent local symptoms. Stent and stent graft with or without the simultaneous use of coil embolization can modulate blood flow and reconstruct the parent artery^32^. Technical success was achieved in all type III dissections with EVT. Carotid endarterectomy with aneurysmectomy was performed in one patient with type III dissections and dissecting pseudoaneurysm thrombosis. In our experience, for young patients with lesions confined to the carotid bifurcation, patients with intolerance to antithrombotic therapy, and patients with large dissecting pseudoaneurysms with or without thrombosis, open surgical treatment is recommended, rather than EVT. Except for one case of distal stent new entry, no complications or adverse events occurred during the hospitalization period or follow-up period after endovascular or surgical treatment for type III sCAD.

Because of the acute occlusion and thrombosis of the carotid artery, type IV dissections were associated with a high rate of stroke, and the symptoms of stroke were significantly more severe than type I, type II, and type III dissections (*p*<0.05). The occlusion site of type IVA dissections was limited to the extracranial carotid artery while type IVB dissections were associated with occlusion of the intracranial large vessel as well. Owing to the intact functionality of the circle of Willis, type IVA dissections usually have an excellent prognosis with medical treatment. In our study, all patients with type IVA dissections had improved symptoms after medical therapy and no ipsilateral recurrent stroke occurred. Only one patient received CAS after the acute phase. Ischemic stroke caused by type IVB dissections, due to the presence of tandem occlusion, if not treated in time, can commonly lead to poor prognosis and severe disability. In our experience, although the difference was not statistically significant due to limited sample size, type IVB dissections were more likely to receive acute phase intervention due to distal embolization or symptoms deterioration after medical treatment than type IVA dissections. In addition, the hospitalization length was significantly higher in type IVB dissections than in type IVA dissections (13.4±7.9 d versus 8.6±2.1 d, *p*<0.05). The management strategy of type IVB dissections remains insufficiently studied. Thrombolytic and antithrombotic therapy had limited efficacy in type IVB dissections due to the presence of tandem occlusion^33, 34^. Although the level of evidence is low, accumulating evidence supports the efficacy and safety of EVT of type IVB dissections^6, 34-39^. The ESO guidelines recommend mechanical thrombectomy for type IVB dissections with ischemic stroke based on limited evidence^23^. In the present study, one-third of patients with type IVB dissections received acute mechanical thrombectomy and stenting for distal embolization or worsening stroke symptoms. Two patients had SICH after acute revascularization, and one died from secondary cerebral hernia. SICH only occurred in patients who received bridging therapy. This is consistent with the results from the SWISS registry and one meta-analysis reporting a higher risk of hemorrhagic complications in patients receiving bridging therapy^40, 41^. Stent occlusion occurred in two IVB dissections after stent implantation. Despite the results, we believe EVT is still required in case of flow limiting dissection in type IVB dissections^6, 23, 34, 35^.

### Limitation

Our study has several limitations. First, since it was a single-center, non-randomized, retrospective observational study, in which selection bias and other confounding factors were inherent. Second, the choice of treatment was left to the discretion of each treating physician, and even within each group, there was some variation in the use of endovascular therapy and medical management. Third, the follow-up period was relatively short. The natural history of sCAD and the prognosis after EVT should be monitored over a longer period, which is underway. Prospective randomized clinical trials with larger sample sizes would be optimal, but the feasibility and practical nature of such task is debatable.

## Conclusions

This study proposes a new and more comprehensive classification method for sCAD, with implication in its medical and surgical management. Antithrombotic therapy is beneficial to reduce the risk of recurrent stroke for stable sCAD. Post-acute phase EVT can be an alternative therapeutic approach for sCAD patients presenting with recurrent symptoms, flow-limiting dissection (type II, type III, and type IVA), or enlarging dissecting pseudoaneurysm (type III). The combination of urgent mechanical thrombectomy and stenting is an effective treatment strategy to treat type IVB dissections with aggravation of symptoms. The short-term results of EVT are encouraging, but the long-term outcome still need to be elucidated.

## Data Availability

The datasets used or analysed during the current study are available from the corresponding author on reasonable request.

## Disclosures

1. Bogousslavsky J, Despland PA, Regli F. Spontaneous carotid dissection with acute stroke. Archives of neurology. 1987;44(2):137–40.

2. Hynes N, Kavanagh EP, Sultan S, Jordan F. Surgical and radiological interventions for treating symptomatic extracranial cervical artery dissection. The Cochrane database of systematic reviews. 2021;2:Cd013118.

3. Debette S, Leys D. Cervical-artery dissections: predisposing factors, diagnosis, and outcome. The Lancet Neurology. 2009;8(7):668–78.

4. Keser Z, Meschia JF, Lanzino G. Craniocervical Artery Dissections: A Concise Review for Clinicians. Mayo Clinic proceedings. 2022;97(4):777–83.

5. Jensen J, Salottolo K, Frei D, Loy D, McCarthy K, Wagner J, et al. Comprehensive analysis of intra-arterial treatment for acute ischemic stroke due to cervical artery dissection. Journal of neurointerventional surgery. 2017;9(7):654–8.

6. Bernardo F, Nannoni S, Strambo D, Puccinelli F, Saliou G, Michel P, et al. Efficacy and safety of endovascular treatment in acute ischemic stroke due to cervical artery dissection: A 15-year consecutive case series. International journal of stroke : official journal of the International Stroke Society. 2019;14(4):381–9.

7. Berge E, Whiteley W, Audebert H, De Marchis GM, Fonseca AC, Padiglioni C, et al. European Stroke Organisation (ESO) guidelines on intravenous thrombolysis for acute ischaemic stroke. European stroke journal. 2021;6(1):I–lxii.

8. Ricotta JJ, Aburahma A, Ascher E, Eskandari M, Faries P, Lal BK. Updated Society for Vascular Surgery guidelines for management of extracranial carotid disease: executive summary. Journal of vascular surgery. 2011;54(3):832–6.

9. Martinelli O, Venosi S, BenHamida J, Malaj A, Belli C, Irace FG, et al. Therapeutical Options in the Management of Carotid Dissection. Annals of vascular surgery. 2017;41:69–76.

10. Chen Z, Chen L, Zhang J, Chen Y, Liu C, Diao Y, et al. Management of Extracranial Carotid Artery Aneurysms: A 6-Year Case Series. Medical science monitor : international medical journal of experimental and clinical research. 2019;25:4933–40.

11. Bontinis V, Antonopoulos CN, Bontinis A, Koutsoumpelis A, Zymvragoudakis V, Rafailidis V, et al. A Systematic Review and Meta-Analysis of Carotid Artery Stenting for the Treatment of Cervical Carotid Artery Dissection. European journal of vascular and endovascular surgery : the official journal of the European Society for Vascular Surgery. 2022;64(4):299–308.

12. Rotzinger DC, Mosimann PJ, Meuli RA, Maeder P, Michel P. Site and Rate of Occlusive Disease in Cervicocerebral Arteries: A CT Angiography Study of 2209 Patients with Acute Ischemic Stroke. AJNR American journal of neuroradiology. 2017;38(5):868–74.

13. Zhang L, Liu X, Gong B, Li Q, Luo T, Lv F, et al. Increased Internal Carotid Artery Tortuosity is a Risk Factor for Spontaneous Cervicocerebral Artery Dissection. European journal of vascular and endovascular surgery : the official journal of the European Society for Vascular Surgery. 2021;61(4):542–9.

14. Putaala J, Metso AJ, Metso TM, Konkola N, Kraemer Y, Haapaniemi E, et al. Analysis of 1008 consecutive patients aged 15 to 49 with first-ever ischemic stroke: the Helsinki young stroke registry. Stroke. 2009;40(4):1195–203.

15. Perry BC, Al-Ali F. Spontaneous cervical artery dissection: the borgess classification. Frontiers in neurology. 2013;4:133.

16. Patel RR, Adam R, Maldjian C, Lincoln CM, Yuen A, Arneja A. Cervical carotid artery dissection: current review of diagnosis and treatment. Cardiology in review. 2012;20(3):145–52.

17. Pelkonen O, Tikkakoski T, Leinonen S, Pyhtinen J, Lepojärvi M, Sotaniemi K. Extracranial internal carotid and vertebral artery dissections: angiographic spectrum, course and prognosis. Neuroradiology. 2003;45(2):71–7.

18. Paraskevas KI, Batchelder AJ, Naylor AR. Fate of Distal False Aneurysms Complicating Internal Carotid Artery Dissection: A Systematic Review. European journal of vascular and endovascular surgery : the official journal of the European Society for Vascular Surgery. 2016;52(3):281–6.

19. Larsson SC, King A, Madigan J, Levi C, Norris JW, Markus HS. Prognosis of carotid dissecting aneurysms: Results from CADISS and a systematic review. Neurology. 2017;88(7):646–52.

20. Brown SC, Falcone GJ, Hebert RM, Yaghi S, Mac Grory B, Stretz C. Stenting for Acute Carotid Artery Dissection. Stroke. 2020;51(1):e3–e6.

21. Morel A, Naggara O, Touzé E, Raymond J, Mas JL, Meder JF, et al. Mechanism of ischemic infarct in spontaneous cervical artery dissection. Stroke. 2012;43(5):1354–61.

22. Blum CA, Yaghi S. Cervical Artery Dissection: A Review of the Epidemiology, Pathophysiology, Treatment, and Outcome. Archives of neuroscience. 2015;2(4).

23. Debette S, Mazighi M, Bijlenga P, Pezzini A, Koga M, Bersano A, et al. ESO guideline for the management of extracranial and intracranial artery dissection. European stroke journal. 2021;6(3):Xxxix–lxxxviii.

24. Kernan WN, Ovbiagele B, Black HR, Bravata DM, Chimowitz MI, Ezekowitz MD, et al. Guidelines for the prevention of stroke in patients with stroke and transient ischemic attack: a guideline for healthcare professionals from the American Heart Association/American Stroke Association. Stroke. 2014;45(7):2160–236.

25. Markus HS, Levi C, King A, Madigan J, Norris J, Cervical Artery Dissection in Stroke Study I. Antiplatelet Therapy vs Anticoagulation Therapy in Cervical Artery Dissection: The Cervical Artery Dissection in Stroke Study (CADISS) Randomized Clinical Trial Final Results. JAMA Neurol. 2019;76(6):657–64.

26. Engelter ST, Traenka C, Gensicke H, Schaedelin SA, Luft AR, Simonetti BG, et al. Aspirin versus anticoagulation in cervical artery dissection (TREAT-CAD): an open-label, randomised, non-inferiority trial. The Lancet Neurology. 2021;20(5):341–50.

27. Brott TG, Halperin JL, Abbara S, Bacharach JM, Barr JD, Bush RL, et al. 2011 ASA/ACCF/AHA/AANN/AANS/ACR/ASNR/CNS/SAIP/SCAI/SIR/SNIS/SVM/SVS guideline on the management of patients with extracranial carotid and vertebral artery disease: executive summary: a report of the American College of Cardiology Foundation/American Heart Association Task Force on Practice Guidelines, and the American Stroke Association, American Association of Neuroscience Nurses, American Association of Neurological Surgeons, American College of Radiology, American Society of Neuroradiology, Congress of Neurological Surgeons, Society of Atherosclerosis Imaging and Prevention, Society for Cardiovascular Angiography and Interventions, Society of Interventional Radiology, Society of NeuroInterventional Surgery, Society for Vascular Medicine, and Society for Vascular Surgery. Journal of the American College of Cardiology. 2011;57(8):1002–44.

28. Moon K, Albuquerque FC, Cole T, Gross BA, McDougall CG. Stroke prevention by endovascular treatment of carotid and vertebral artery dissections. Journal of neurointerventional surgery. 2017;9(10):952–7.

29. Wang G, Li C, Piao J, Xu B, Yu J. Endovascular treatment of blunt injury of the extracranial internal carotid artery: the prospect and dilemma. International journal of medical sciences. 2021;18(4):944–52.

30. Asif KS, Lazzaro MA, Teleb MS, Fitzsimmons BF, Lynch J, Zaidat O. Endovascular reconstruction for progressively worsening carotid artery dissection. Journal of neurointerventional surgery. 2015;7(1):32–9.

31. Seven NA, Casanegra AI, Lanzino G, Keser Z. Extracranial Internal Carotid and Vertebral Dissecting Pseudoaneurysms: Clinical Features and Long-Term Outcomes. Stroke: Vascular and Interventional Neurology. 2023;3(3).

32. Akinduro OO, Gopal N, Hasan TF, Nourollah-Zadeh E, Vakharia K, De Leacy R, et al. Pipeline Embolization Device for Treatment of Extracranial Internal Carotid Artery Pseudoaneurysms: A Multicenter Evaluation of Safety and Efficacy. Neurosurgery. 2020;87(4):770–8.

33. Rubiera M, Ribo M, Delgado-Mederos R, Santamarina E, Delgado P, Montaner J, et al. Tandem internal carotid artery/middle cerebral artery occlusion: an independent predictor of poor outcome after systemic thrombolysis. Stroke. 2006;37(9):2301–5.

34. Papanagiotou P, Haussen DC, Turjman F, Labreuche J, Piotin M, Kastrup A, et al. Carotid Stenting With Antithrombotic Agents and Intracranial Thrombectomy Leads to the Highest Recanalization Rate in Patients With Acute Stroke With Tandem Lesions. JACC Cardiovascular interventions. 2018;11(13):1290–9.

35. Marnat G, Bühlmann M, Eker OF, Gralla J, Machi P, Fischer U, et al. Multicentric Experience in Distal-to-Proximal Revascularization of Tandem Occlusion Stroke Related to Internal Carotid Artery Dissection. AJNR American journal of neuroradiology. 2018;39(6):1093–9.

36. Turc G, Bhogal P, Fischer U, Khatri P, Lobotesis K, Mazighi M, et al. European Stroke Organisation (ESO) -European Society for Minimally Invasive Neurological Therapy (ESMINT) Guidelines on Mechanical Thrombectomy in Acute Ischaemic StrokeEndorsed by Stroke Alliance for Europe (SAFE). European stroke journal. 2019;4(1):6–12.

37. Da Ros V, Scaggiante J, Pitocchi F, Sallustio F, Lattanzi S, Umana GE, et al. Mechanical thrombectomy in acute ischemic stroke with tandem occlusions: impact of extracranial carotid lesion etiology on endovascular management and outcome. Neurosurgical focus. 2021;51(1):E6.

38. Karam A, Bricout N, Khyeng M, Cordonnier C, Leclerc X, Henon H, et al. Safety and outcome of mechanical thrombectomy in ischaemic stroke related to carotid artery dissection. Journal of neurology. 2022;269(2):772–9.

39. Li S, Zi W, Chen J, Zhang S, Bai Y, Guo Y, et al. Feasibility of Thrombectomy in Treating Acute Ischemic Stroke Because of Cervical Artery Dissection. Stroke. 2018;49(12):3075–7.

40. Hao Y, Zhang Z, Zhang H, Xu L, Ye Z, Dai Q, et al. Risk of Intracranial Hemorrhage after Endovascular Treatment for Acute Ischemic Stroke: Systematic Review and Meta-Analysis. Interventional neurology. 2017;6(1-2):57–64.

41. Traenka C, Jung S, Gralla J, Kurmann R, Stippich C, Simonetti BG, et al. Endovascular therapy versus intravenous thrombolysis in cervical artery dissection ischemic stroke - Results from the SWISS registry. European stroke journal. 2018;3(1):47–56.

